# PROSPECTIVE OBSERVATIONAL STUDY ON THE ASSOCIATION BETWEEN PERIOPERATIVE CORTISOL LEVEL AND HEMODYNAMIC VARIABLES IN PATIENTS UNDERGOING PITUITARY SURGERIES

**DOI:** 10.64898/2025.12.30.25343221

**Authors:** Praveen R Sarguru, Unnikrishnan Prathapadas, Smita Vimala, Prakash Nair, Manikandan Sethuraman, Ajay Prasad Hrishi, Ranganatha Praveen C S, Bijith Vishnu

## Abstract

**Background:** Although transnasal transsphenoidal (TNTS) resection is the standard treatment for pituitary adenomas, the procedure carries a risk of hypothalamic-pituitary-adrenal (HPA) axis disruption. This potential for hypocortisolism often prompts the empiric use of perioperative steroids to mitigate hemodynamic instability. However, there is a paucity of data strictly correlating serum cortisol concentrations with intraoperative blood pressure fluctuations. This study sought to evaluate the relationship between perioperative cortisol levels, hemodynamic stability, and the necessity for vasopressors, while also examining the efficacy of prophylactic steroid administration.

**Methods:** We conducted a prospective observational study involving 90 adult patients (aged 18–60) undergoing elective TNTS at a tertiary care center. Serial serum cortisol measurements were taken at baseline, on the morning of surgery, post-induction, post-sphenid drilling, and during episodes of refractory hypotension. Continuous hemodynamic monitoring included heart rate and arterial pressures (systolic, diastolic, and mean). Hydrocortisone prophylaxis was administered to high-risk patients or those with baseline cortisol <80 ng/ml. Statistical associations were determined using Spearman correlation and multivariate logistic regression models.

**Results:** Our analysis revealed no statistically significant association between perioperative cortisol levels and the incidence of hypotension or vasopressor requirements across any time point. Furthermore, the administration of prophylactic hydrocortisone failed to demonstrate a reduction in the occurrence of intraoperative hemodynamic instability.

---

As the second most common benign intracranial tumor, pituitary adenomas account for roughly 10–15% of such cases. They are predominantly managed via endoscopic transnasal transsphenoidal (TNTS) surgery, an approach that offers reduced morbidity and faster recovery times by eliminating the need for brain retraction. However, the management of these patients is unique due to the inherent risks associated with HPA axis dysfunction and subsequent hemodynamic volatility.

Cortisol plays a critical role in the body’s stress response. In patients with pituitary disease, however, the tumor may impair the HPA axis either through direct mass effect on the gland or via hormonal interference. (1) While surgery is a controlled stressor that requires an adequate cortisol response to preserve blood pressure and metabolic function, these patients often lack the reserve to mount such a response. (2)

Historically, the fear of intraoperative hypotension has driven the routine use of perioperative stress-dose steroids. (3) Yet, the necessity of this universal approach is currently being debated. Although steroids protect against adrenal insufficiency, their use is not without consequence; they carry risks of significant complications such as hyperglycemia, immunosuppression, and issues with wound repair.

Data regarding perioperative hemodynamics in pituitary surgery remains sparse. Previous investigations have largely confined their analysis to static biochemical profiles or downstream outcomes like diabetes insipidus. Few studies have attempted to correlate real-time cortisol dynamics with intraoperative cardiovascular stability. Although Guo et al. (2022) and Sterl et al. (2019) investigated the feasibility of withholding steroids, these studies lacked a granular assessment of intraoperative hemodynamics. (4,5) Likewise, Lee et al. (2021) compared hydrocortisone against placebo without assessing the relationship between serial cortisol levels and vasopressor requirements.

Therefore, a critical question remains unanswered: Do perioperative cortisol levels directly influence intraoperative blood pressure variability and the necessity for vasopressor support? Answering this is essential to determine whether the current paradigm of routine stress-dose supplementation should be replaced by a personalized management strategy.

## METHODS

### Study Design and Setting

This prospective observational investigation was undertaken between July 2023 and April 2025 at the Sree Chitra Tirunal Institute for Medical Sciences and Technology, Trivandrum. The study adhered to ethical standards with approval from the Institutional Ethics Committee (SEC/IEC/2055/May/2023) and registration under the Clinical Trials Registry of India (CTRI/2023/06/053493).

### Eligibility Criteria

- **Inclusion:** Adults (18–60 years) with ASA status I–III undergoing elective endoscopic TNTS pituitary surgery with a preoperative GCS of 15.
- **Exclusion:** We excluded patients with Cushing’s disease, documented prior adrenal insufficiency/surgery, or those receiving long-term steroid therapy. Further exclusions included ASA class IV–V, pre-existing hypertension, emergency surgical cases, pregnancy, lactation, or refusal to consent.

### Methods

Potential candidates were identified from the elective neurosurgical operating schedule. During the pre-anesthetic evaluation, we recorded baseline demographic data, hormonal profiles including cortisol(SC baseline), lesion characteristics, and vital signs, including heart rate and blood pressure. On the morning of surgery, an initial serum cortisol sample (SC1) was drawn at 7:30 AM.

A standardized anesthetic protocol was strictly enforced. Induction was achieved using fentanyl 2µg/kg and propofol 2-3 mg/kg. Following mask ventilation, neuromuscular blockade was established with atracurium 0.5 mg/kg to facilitate intubation. Maintenance anesthesia consisted of sevoflurane (MAC > 0.7) in an oxygen/air mixture (FiO2 0.5), alongside continuous infusions of atracurium 10µg/kg/min and fentanyl 2µg/kg/hour.Targeted BIS monitoring (40–60) continued throughout the procedure.

Regarding steroid supplementation, our institutional protocol mandated hydrocortisone 100mg at induction only for specific high-risk groups: patients with baseline serum cortisol <80ng/ml, hyponatremia (Na< 135mEq/L), or coronary artery disease.

Serial serum cortisol measurements were timed to capture physiological stress responses: immediately post-induction (SC2) and following sphenoid drilling (SC3). An additional conditional sample (SCx) was reserved for episodes of refractory hypotension.

Hemodynamic stability was tracked continuously using a Philips IntelliVue MX700 monitor. Hypotension was defined as systolic blood pressure (SBP) <100mmHg, mean arterial pressure (MAP)<65mmHg or a decline of 20% from baseline. Refractory hypotension was characterized as hypotension unresponsive to fluid resuscitation requiring vasopressor escalation for >3 minutes. Total vasopressor consumption (mephentermine and noradrenaline) was quantified at the procedure’s conclusion.

### Sample Size Calculation

Utilizing G*Power software, we calculated the requisite sample size based on a projected correlation coefficient ($r$) of 0.3. With parameters set to an alpha of 0.05 and a power of 0.8, the minimum cohort size was established at 85. Consequently, 90 patients were enrolled to offset potential exclusions or dropouts.

### Statistical Methodology

Data analysis was executed using SPSS v25.

- **Descriptive Statistics:** Continuous data are expressed as mean $\pm$ SD or median (IQR) based on normality testing; categorical data are reported as counts and percentages.
- **Correlation & Regression:** We assessed the association between cortisol levels and hypotensive episodes using Spearman correlation. To evaluate predictors of hypotension—specifically cortisol levels and steroid use—we applied logistic regression analysis.
- **Multivariate Analysis:** Confounders were controlled for using multivariate techniques.
- **Significance:** A $p\text{-value} \le 0.05$ was considered statistically significant.

## Results

### Patient Characteristics

Out of 124 screened patients, 3 patients who refused to participate, 4 patients with Cushing’s disease and 27 with hypertension were excluded. 90 patients met inclusion criteria; 54 females (60%) and 36 males (40%), median age 50 years (IQR 41.5–58.5). ASA classification included 11 (12.22%) Grade I, 38 (42.22%) Grade II, and 41 (45.56%) Grade III. Pituitary adenoma types were predominantly non-functioning tumors (65.56%) with 34.44% functioning tumors.

**Table 1.** Baseline characteristics.

| Variable | Value |
| --- | --- |
| Age, median (IQR), years | 50 (41.5–58.5) |
| Gender, n (%) | Female: 54 (60);<br>Male: 36 (40) |
| ASA Grade, n (%) | I: 11 (12.2); |
|  | II: 38 (42.2);<br>III: 41 (45.6) |
| Tumor type, n (%) | Non-functioning: 59 (65.6);<br>Functioning: 31 (34.4) |
| Recommended Preinduction hydrocortisone, n (%) | Yes: 49 (54.4); No: 41 (45.6) |
| Duration of surgery, minutes, median (IQR) | 245 (210–290) |

### Perioperative Cortisol Dynamics

Based on institutional protocol, 49 patients (54.4%) were recommended to receive pre-induction hydrocortisone, while 41 patients (45.6%) were not. As expected, the group recommended for hydrocortisone had a significantly lower median baseline serum cortisol level compared to the group not receiving steroids (87.5 ng/mL vs. 137 ng/mL, p < 0.001). Following administration, the hydrocortisone group exhibited supraphysiological cortisol levels post-induction (median 616 ng/mL) and after sphenoid drilling (median 492 ng/mL), which were significantly higher than in the non-steroid group (p < 0.001 for both). Median cortisol values showed expected circadian rise at SC1. Subsequent levels (SC2, SC3) did not significantly differ between steroid-supplemented and non-supplemented groups.

### Hemodynamic Correlations

#### Primary Outcome

Correlation Between Serum Cortisol and Intraoperative Hypotension.

No significant correlation was found between perioperative serum cortisol levels and the incidence of intraoperative hypotension, irrespective of the definition used.

#### SBP < 100 mmHg

Spearman correlation analysis between cortisol levels at all measured time points (baseline, 7:30 AM, post-induction, post-sphenoid drilling) and the number of hypotensive episodes showed weak, non-significant correlations (correlation coefficients ranged from -0.107 to 0.061; all p > 0.05).

#### MAP < 65 mmHg

Similarly, no significant correlation was observed between cortisol levels and hypotensive episodes defined by a MAP < 65 mmHg (correlation coefficients ranged from -0.165 to 0.12; all p > 0.05).

#### ≥20% Drop in MAP

The analysis also revealed no significant correlation when hypotension was defined as a ≥20% decrease from baseline MAP (correlation coefficients ranged from -0.09 to +0.148; all p > 0.05).

Logistic regression analysis confirmed these findings. Neither baseline serum cortisol nor the 7:30 AM cortisol level were significant predictors of intraoperative hypotension. Furthermore, multivariate analysis demonstrated that pre-induction hydrocortisone administration was not associated with a reduced risk of hypotension under any of the three definitions.

#### Secondary Outcomes

Vasopressor Requirement and Hemodynamic Stability The administration of pre-induction hydrocortisone did not significantly affect vasopressor requirements or overall hemodynamic stability.

The median mephentermine requirement was comparable between the group that received hydrocortisone (12 mg; IQR, 0-18) and the group that did not (9 mg; IQR, 0-18), with no statistically significant difference (p = 0.974).

Refractory hypotension requiring noradrenaline infusion occurred in only three patients. All three of these patients were in the group that had been recommended for and had received pre-induction hydrocortisone. No instances of refractory hypotension occurred in the group that did not receive steroids.

**Table 2.** Vasopressor use by hydrocortisone status.

| Variable | Hydrocortisone<br>(n=49) | No Hydrocortisone<br>(n=41) | pvalue |
| --- | --- | --- | --- |
| Mephentermine dose, mg,<br>median (IQR) | 15 (10–25) | 14 (10–23) | 0.74 |
| Noradrenaline dose, µg,<br>mean ± SD | 112 ± 30 | 108 ± 28 | 0.62 |

## Discussion

### Comparison with Previous Literature

Our results are consistent with multiple contemporary studies:

Sterl et al. (2019): Found that patients without steroids had higher postoperative morning cortisol but no difference in intraoperative hemodynamic outcomes.(4)

Lee et al. (2021): Double-blind trial found no difference in intraoperative blood pressure or heart rate with hydrocortisone versus placebo.(1)

Guo et al. (2022): Demonstrated withholding hydrocortisone was as safe as administering it in patients with intact HPA axis, with fewer metabolic complications.(5)

By integrating serial cortisol measurements and continuous hemodynamic monitoring, our study adds granularity missing from these earlier works.

This prospective observational study fundamentally challenges the long-held clinical paradigm that perioperative serum cortisol levels are a primary determinant of intraoperative hemodynamic stability in patients undergoing pituitary surgery. Our principal finding is a consistent and robust lack of a statistically significant association between serum cortisol concentrations and the incidence of intraoperative hypotension or the requirement for vasopressor support. This conclusion remained steadfast across three distinct, clinically relevant definitions of hypotension, underscoring its validity. Furthermore, the prophylactic administration of a conventional “stress dose” of hydrocortisone did not confer any observable benefit in maintaining hemodynamic stability or reducing vasopressor needs, suggesting that this routine practice may not be necessary for intraoperative management.

The practice of administering routine perioperative steroids is deeply entrenched, rooted in historical case reports from the 1950s that documented fatal outcomes from acute adrenal insufficiency following surgery. This history fostered a clinical dogma favoring universal steroid coverage, based on the well-understood physiological role of cortisol in maintaining vascular tone and sensitizing adrenergic receptors to catecholamines. However, our findings suggest a more intricate reality within the modern, pharmacologically sophisticated anesthetic environment. The direct, linear relationship between a patient’s serum cortisol and their moment-to-moment blood pressure appears to be absent. In our cohort, patients who did not receive hydrocortisone demonstrated a preserved endogenous stress response, with cortisol levels rising appropriately in response to surgical stimuli, indicating a functionally intact hypothalamic-pituitary-adrenal (HPA) axis. Conversely, patients who received prophylactic hydrocortisone based on a low baseline reading were driven into a state of profound supraphysiological hypercortisolemia. This striking observation reframes the central clinical question from “How do we treat low cortisol?” to “Are we causing unnecessary and extreme hypercortisolemia with routine prophylaxis?”

Our results are consistent with a growing body of literature that questions the necessity of universal steroid supplementation. A randomized controlled trial by Lee et al. found no significant difference in intraoperative hypotension between patients receiving preoperative hydrocortisone versus placebo. Similarly, a retrospective study by Regan et al., which employed a selective steroid protocol based on a morning cortisol threshold, reported no instances of cardiovascular collapse or symptomatic hypotension despite the majority of patients not receiving empirical steroids. Our findings also align with the work of Alexander et al. and Guo et al., who found that a selective approach to steroid administration did not increase postoperative complications or the incidence of adrenal insufficiency. While these studies focused primarily on the safety of withholding steroids and postoperative outcomes, our study provides the missing intraoperative physiological data. By directly correlating real-time cortisol levels with continuous hemodynamic monitoring, we offer a mechanistic insight that supports their conclusions: variations in perioperative cortisol do not appear to translate into acute hemodynamic compromise. The convergence of these studies, despite methodological differences, strengthens the evidence against a direct relationship between perioperative cortisol fluctuations and acute hemodynamic outcomes.

The clinical implications of our data are significant. The fear of cortisol-dependent hypotension has driven the routine use of stress-dose steroids, exposing a large number of patients to the welldocumented risks of supraphysiological glucocorticoids, including hyperglycemia, impaired wound healing, increased infection risk, and the potential masking of true postoperative adrenal insufficiency. This study provides a robust, evidence-based argument that this fear may be largely unfounded in the modern operating room, where advanced monitoring and potent, rapid-acting vasopressors allow for the immediate and effective management of hemodynamic instability, irrespective of its cause. Our findings empower clinicians to move away from a “one-size-fits-all” protocol towards a more judicious, selective, and individualized approach, guided by preoperative endocrine assessment and vigilant intraoperative monitoring.

### Limitations

- Single-center study; generalizability may be limited.
- Exclusion of hypertensive and Cushing’s patients reduces applicability to broader populations.
- Incidence of refractory hypotension was low (3 cases), limiting conclusions on this subgroup.
- Observational design precludes causal inference.

### Future Directions

Multicenter randomized controlled trials should further investigate:

- Cortisol thresholds for supplementation.
- Outcomes in higher-risk cohorts (Cushing’s disease, elderly, hypertensives).
- Long-term effects of withholding steroids on endocrine recovery and quality of life.

### Conclusion

Our prospective observational study shows that:

- Perioperative cortisol levels are not significantly correlated with intraoperative hypotension or vasopressor use in pituitary surgery.
- Hydrocortisone supplementation did not reduce hypotension incidence or vasopressor requirement.
- Incidence of refractory hypotension was rare, and its occurrence was not mitigated by steroid administration.

These findings strongly support a more selective, individualized approach to perioperative steroid use rather than routine universal supplementation. Avoiding unnecessary hydrocortisone reduces risks of metabolic and infectious complications while maintaining hemodynamic safety. Our work contributes to the evolving paradigm that routine stress-dose steroids are not warranted in all patients undergoing pituitary surgery and reinforces the need for further randomized studies to refine guidelines.

## Data Availability

All data produced in the present work are contained in the manuscript

## REFERENCES

1. Lee HC, Yoon HK, Kim JH, Kim YH, Park HP. Comparison of intraoperative cortisol levels after preoperative hydrocortisone administration versus placebo in patients without adrenal insufficiency undergoing endoscopic transsphenoidal removal of nonfunctioning pituitary adenomas: a double-blind randomized trial. 2020 Jan 24 [cited 2025 July 7]; Available from: https://thejns.org/view/journals/j-neurosurg/134/2/article-p526.xml

2. Taşar S, Dikmen N, Bulut İ, Haskılıç YE, Saç RÜ, Şenes M, et al. Potential role of salivary cortisol levels to reflect stress response in children undergoing congenital heart surgery. Cardiol Young. 2023 Jan;33(1):106–12.

3. Yeh PJ, Chen JW. Pituitary tumors: surgical and medical management. Surg Oncol. 1997 Aug 1;6(2):67–92.

4. Sterl K, Thompson B, Goss CW, Dacey RG, Rich KM, Zipfel GJ, et al. Withholding Perioperative Steroids in Patients Undergoing Transsphenoidal Resection for Pituitary Disease: Randomized Prospective Clinical Trial to Assess Safety. Neurosurgery. 2019 Aug;85(2):E226.

5. Guo X, Zhang D, Pang H, Wang Z, Gao L, Wang Y, et al. Safety of Withholding Perioperative Hydrocortisone for Patients With Pituitary Adenomas With an Intact Hypothalamus-Pituitary-Adrenal Axis: A Randomized Clinical Trial. JAMA Netw Open. 2022 Nov 16;5(11):e2242221.

